# Racial and Ethnic Differences in the Association between Depressive Symptoms and Cognitive Outcomes in Older Adults: Findings from KHANDLE and STAR

**DOI:** 10.1101/2023.09.07.23295205

**Authors:** Marcia P Jimenez, Emma L. Gause, Eleanor Hayes-Larson, Emily P. Morris, Evan Fletcher, Jennifer Manly, Paola Gilsanz, Yenee Soh, Maria Corrada, Rachel A. Whitmer, M. Maria Glymour

## Abstract

**INTRODUCTION:** Depressive symptoms are associated with higher risk of dementia but how they impact cognition in diverse populations is unclear.

**METHODS:** Asian, Black, LatinX, or White participants (n=2,227) in the Kaiser Healthy Aging and Diverse Life Experiences (age 65+) and the Study of Healthy Aging in African Americans (age 50+) underwent up to three waves of cognitive assessments over four years. Multilevel models stratified by race/ethnicity were used to examine whether depressive symptoms were associated with cognition or cognitive decline and whether associations differed by race/ethnicity.

**RESULTS:** Higher depressive symptoms were associated with lower baseline verbal episodic memory scores (−0.06, 95%CI:-0.12,-0.01; −0.15, 95%CI:-0.25,-0.04), and faster decline annually in semantic memory (−0.04, 95%CI:-0.07,-0.01; −0.10, 95%CI:-0.15,-0.05) for Black and LatinX participants. Depressive symptoms were associated with lower baseline but not decline in executive function.

**DISCUSSION:** Depressive symptoms were associated with worse cognitive domains, with some evidence of heterogeneity across racial/ethnic groups.

**Highlights:**

- We examined whether baseline depressive symptoms were differentially associated with domain-specific cognition or cognitive decline by race/ethnicity.
- Depressive symptoms were associated with worse cognitive scores for all racial/ethnic groups across different domains examined.
- Higher depressive symptoms were associated with faster cognitive decline for semantic memory for Black and LatinX participants.
- The results suggest a particularly harmful association between depressive symptoms and cognition in certain racial/ethnic groups.

## BACKGROUND

As the older adult population grows in the U.S., it will become more racially and ethnically diverse. Given the higher burden of dementia among Black and Latino individuals, uncovering modifiable risk factors of Alzheimer’s Disease and Related Dementias (ADRD) is essential.^1^ Convincing evidence – derived predominantly in samples comprising non-Hispanic Black and White-identified participants - supports the notion that depression increases risk of cognitive decline.^2–8^ Since depression can be treated, it is an attractive modifiable risk factor for the prevention or attenuation of cognitive decline in older adults. However, the relationships between race/ethnicity and depressive symptoms, and between depressive symptoms and cognition are complex.^9,10^

The reserve capacity model (RCM) posits that certain subgroups of the population such as individuals of lower socioeconomic status (SES) are more vulnerable to the negative health effects of depression because increased exposure to chronic stress diminishes reserve capacity.^11^ The Minority Stress Theory (MST) asserts that exposure to racial discrimination and acculturative stress constitutes a hostile psychological environment leading to negative health consequences. In other words, RCM in conjunction with MST suggests that Black and LatinX individuals may or may not experience depression more often than White individuals, but they experience more severe health consequences of depression when it is present. The literature on the prevalence of depressive symptoms across race and ethnicity is mixed, some studies have found the prevalence to be lower among White participants, compared to Black and LatinX participants,^12^ while other studies have found a lower prevalence of depressive symptoms among Black and LatinX participants.^13–15^ Furthermore, studies show that when depression affects Black individuals, it is usually untreated and is more severe and disabling compared with that in White individuals, suggesting that the burden of depressive disorders may be higher among Black than White participants.^13^

Currently, the relation of depressive symptoms with late life cognition across racial/ethnic groups is only partially understood. To our knowledge, this is among the first studies to examine the prevalence of depressive symptoms and investigates its relationship with cognition and cognitive decline in a multi-ethnic older sample followed up to 4 years. We hypothesize that the prevalence of depressive symptoms and its association with cognitive health will differ among race and ethnicity groups.

## METHODS

### Data

This study used data from the Kaiser Healthy Aging and Diverse Life Experiences (KHANDLE) and the Study of Healthy Aging in African Americans (STAR) cohorts. KHANDLE comprises community-dwelling older adults in the San Francisco Bay and Sacramento regions of California who were aged 65 or older as of January 1, 2017 and were long-term members of Kaiser Permanente Northern California. KHANDLE participants were eligible for inclusion if they spoke either English or Spanish and participated in at least one voluntary health checkup (Multiphasic Health Checkup [MHC]) with Kaiser Permanente between 1964-1985. Exclusion criteria included a previous diagnosis of dementia or other neurodegenerative disease (frontotemporal dementia, Lewy body disease, Pick’s disease, Parkinson’s disease with dementia, Huntington’s disease), or a diagnosis related to poor health that might interfere with study participation or study interviews (defined by hospice activity in the past 12 months, history of severe chronic obstructive pulmonary disease in the past 6 months, congestive heart failure hospitalizations in the past 6 months, and history of end stage renal disease or dialysis in the past 12 months). Participants selected for inclusion were randomly sampled from this eligible participant pool within strata of race and ethnicity and educational attainment to achieve balanced numbers of participants who reported Asian, Black, LatinX, and White race and ethnicity with diverse educational backgrounds.

The STAR study is similarly comprised of community-dwelling older adults from the San Francisco Bay area, but enrolled only participants who identified as Black or African American that participated in at least one MHC exam and who were aged 50 or older as of January 1, 2018. The STAR study used the same exclusion as KHANDLE, outlined above. More information about the KHANDLE and STAR cohort can be found in cited materials.^16–19^ At wave 1, which we will hereafter refer to as baseline, KHANDLE had 1712 participants and STAR had 764 participants (N=2389). Participants were excluded from the analysis set if they were missing or refused to answer race and ethnicity data, or if they were missing information on any of the primary model covariates (including age at baseline, sex, education, income, and marital status). 4 participants who self-reported as Native American were excluded due to concerns about small numbers and identifiability. The final analysis cohort was made up of 2227 participants. (Figure S1).

### Exposure

The primary exposure of interest was depressive symptoms reported at baseline KHANDLE and STAR interviews. The KHANDLE cohort used the 4-item Patient-Reported Outcomes Measurement Information System (PROMIS) computer adaptive testing (CAT) v1.0 administered through National Institutes of Health (NIH) Toolbox and the STAR cohort used the 8-item PROMIS-57 administered through DatStat. We followed the depression symptoms scoring manual from PROMIS to standardize the PROMIS scores across forms between the two cohorts. These depressive symptoms scores were transformed into theta scores which were standardized to the US adult population so that a difference in one unit represented a standardized difference in depression symptoms which were centered around zero for the average US adult. Depressive symptom theta scores were treated as a continuous variable.

### Outcomes

Cognitive function was assessed across three waves using the Spanish and English Neuropsychological Assessment Scales (SENAS). Both KHANDLE and STAR assessed cognition using SENAS 3 times between 2017 and 2021. The SENAS includes three cognitive domains (executive function, verbal episodic memory, and semantic memory), and was administered either English or Spanish, with language of administration determined by an algorithm that considered preferred language and everyday language usage in a variety of settings. The SENAS is a battery of cognitive tests that has previously undergone extensive development for valid comparisons of cognitive change across racial and ethnic and linguistically diverse groups.^20^ Briefly, item response theory and confirmatory factor analysis methods were used to construct measures that are psychometrically matched across domains with respect to level of reliability across the ability continuum. The Episodic Memory score is derived from a multi-trial word-list-learning test. The Semantic Memory measure is a composite of highly correlated verbal (object-naming) and nonverbal (picture association) tasks. The Executive Function composite is constructed from component tasks of category fluency, phonemic (letter) fluency, and working memory (digit-span backward, list sorting).

Administration procedures, measure development and psychometric characteristics of the SENAS battery are described in detail elsewhere.^20,21^ Due to the use of visual stimuli in the assessment of semantic memory, we were unable to assess semantic memory during the third waves of KHANDLE and STAR since SENAS was administered by phone during the COVID-19 pandemic. SENAS measures were standardized into z-scores using baseline values for analysis across the two study cohorts prior to restricting to complete cases. These z-scores can be interpreted as standard deviations away from the mean of zero for the average individual in the analysis set at baseline.

### Covariates

Race and ethnicity were self-reported at baseline. Individuals were able to select more than one racial and ethnic categories which were collapsed for these analyses as Latino, Black, Asian, or White using that assignment order. Other baseline socio-demographic information included age (in years at time of assessment), biological sex (female/male), educational attainment (operationalized as college degree or higher vs. no college degree), household income (operationalized as greater than 55k annually vs. less than 55k), and marital status (operationalized as married or living with partner vs. not). Sensitivity analyses included additional covariates that could potentially be mediators: daily socializing, daily physical activity, smoking history, heavy drinking, and self-reported health. More information on these variables is provided in the supplementary material (Methods S1).

### Statistical Analysis

We first descriptively compared the distributions of depressive symptoms across race/ethnicity subgroups (following Ward et al),^22^ as well as the distributions of SENAS cognitive domains across these groups. To assess the association between depressive symptoms and cognition and cognitive decline, we used multilevel linear regression with random intercepts for individuals to separately assess the associations of depressive symptoms with baseline and longitudinal change in each SENAS domain (executive function, verbal episodic memory, and semantic memory).

An interaction term between depressive symptoms and years since baseline was included to test for differences in decline in cognitive domain over time. In an additional sensitivity analysis, we also fit a model with interactions between time and all included covariates to control for confounding on estimates of cognitive decline. To examine race/ethnicity as a potential effect modifier of the association between depressive symptoms and cognitive function and subsequent cognitive decline, we used a two-step approach. First, we included an interaction term between race/ethnicity and depression for each cognitive outcome and use a joint likelihood ratio test to determine whether the model including the interaction term was a better fit to the data compared to the nested model without interaction terms. Second, we evaluated associations in models stratified by race and ethnicity.

Our main results are based on complete case analysis (N=2,227), and we also performed supplementary analysis with multiple imputation using chained equations (MICE) on the sample with missing covariates (N=2,469) to impute the most likely missing values. We imputed 10 datasets using all model variables in addition to study cohort, daily socializing, daily physical activity, smoking history, heavy drinking, and self-reported health all measured at baseline, as well as practice setting where the interview took place in each wave. Models were fit to each of the 10 imputed datasets and then results were pooled using Rubin’s Rules. We also performed another sensitivity analysis additionally adjusting for the lifestyle covariates described above as well as the practice setting. Data analyses were conducted using R software version 4.1.3.^23^

## RESULTS

Among the 2,227 individuals included in the analysis, 17% (n=380) were classified as Asian, 48% (n=1076) Black, 14% (n=318) Latino, and 20% (n=453) White (Table 1). The average age of the cohort at baseline was 72.7 years (SD=8.0) and the majority were female (62%).

**Table 1.**
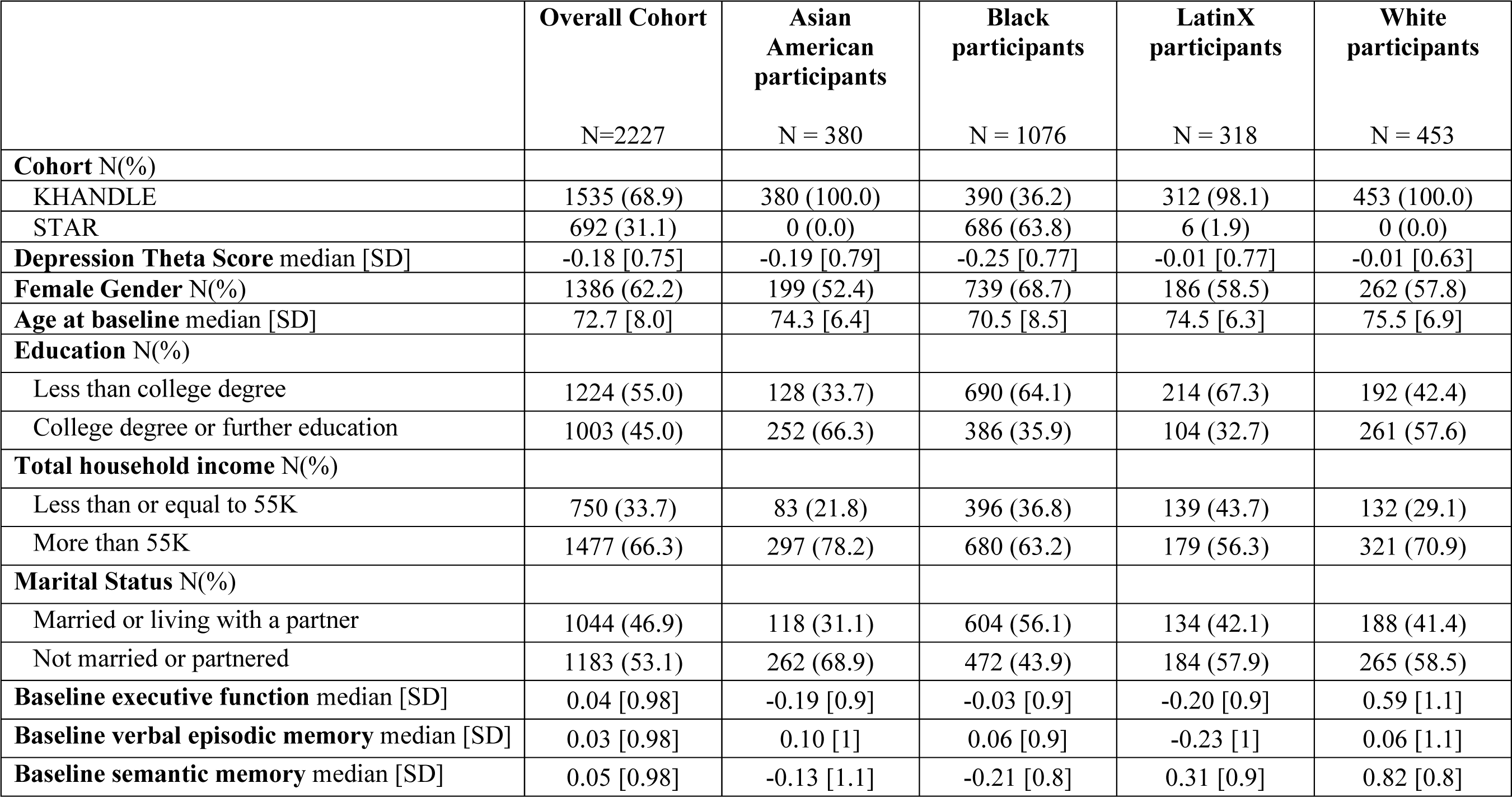
Characteristics of complete case participants in KHANDLE and STAR by race and ethnicity at baseline (N= 2227)

Education, income, and other sociodemographic factors varied across race and ethnicity (Table 1). Depressive symptoms levels were somewhat lower among Asian and Black participants, compared to Latino and White participants (Figure 1).

**Figure 1:**
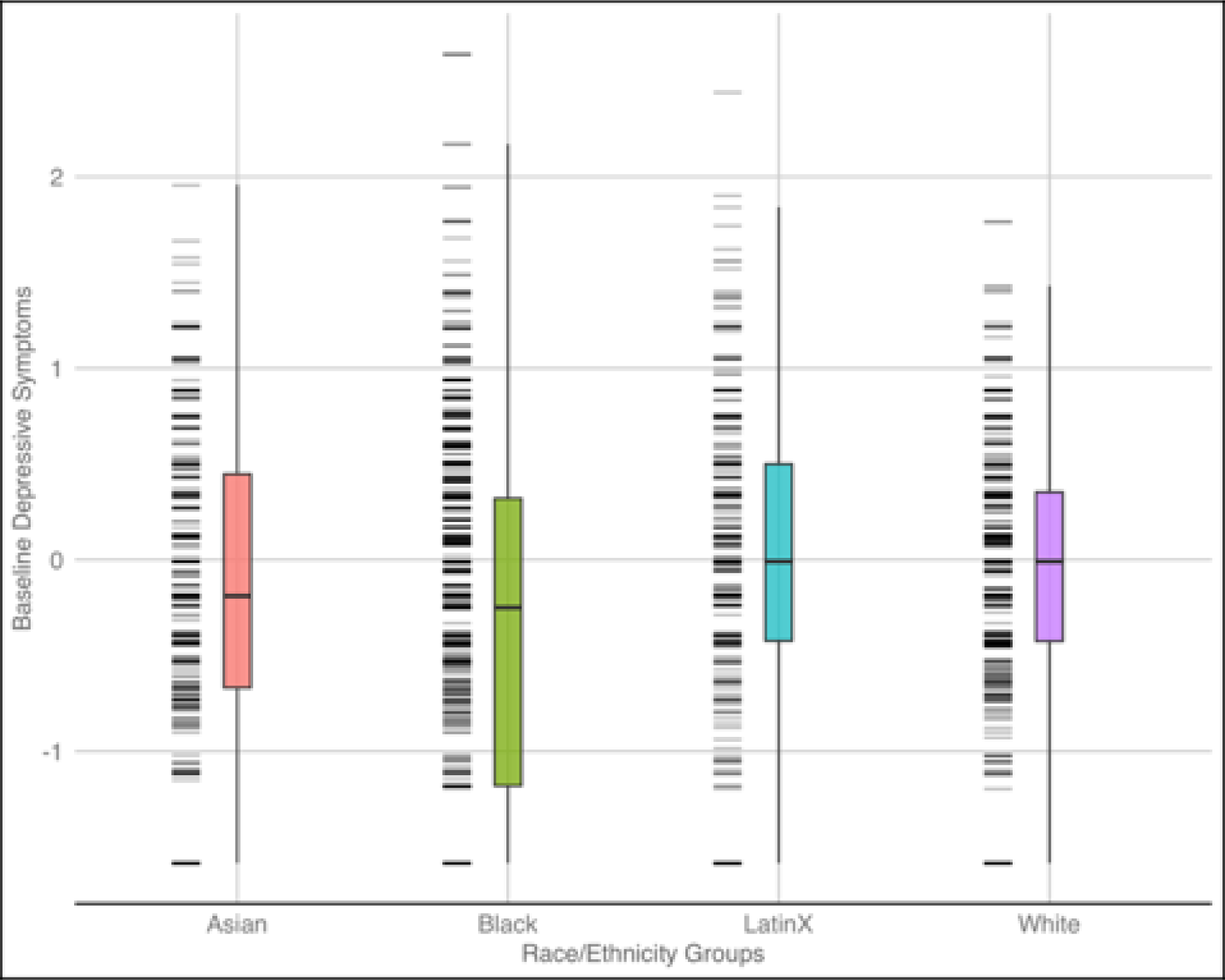
Distribution of baseline depressive symptoms by race and ethnicity groups in KHANDLE and STAR (N= 2227)

### Baseline Cognitive Function

Table 2 shows the estimates for the association of depressive symptoms with baseline cognitive function for the full sample and stratified by race and ethnicity. In the full sample, higher depressive symptoms were associated with −0.06 (95%CI: −0.10, −0.02) SD units lower executive function and −0.05 (95%CI: −0.09, −0.00) SD units lower verbal episodic memory. We did not observe an association between depressive symptoms and semantic memory score in the full sample. The likelihood ratio test suggested a better goodness of fit when interaction terms between depression and race/ethnicity group were included. Stratified analysis by race and ethnicity suggested that higher depressive symptoms at baseline were associated with lower executive function and verbal episodic memory scores among Black and LatinX participants (e.g., −0.06 SD, 95%CI: −0.12, −0.01; −0.17 SD, 95%CI: −0.28, −0.06 for executive function for Black and Latino participants respectively). Higher depressive symptoms at baseline were associated with lower semantic memory scores among LatinX participants (−0.15, 95%CI:-0.27, - 0.04), but not among Asian, Black or White participants. Results of the sensitivity analysis using the multiply imputed data showed similar associations between baseline depressive symptoms and lower scores on specific cognitive domains. For example, in the full sample, higher depressive symptoms were associated with −0.07 (95%CI: −0.11, −0.03) SD units lower executive function and −0.06 (95%CI: −0.10, −0.02) SD units lower verbal episodic memory (Table S1).

**Table 2.**
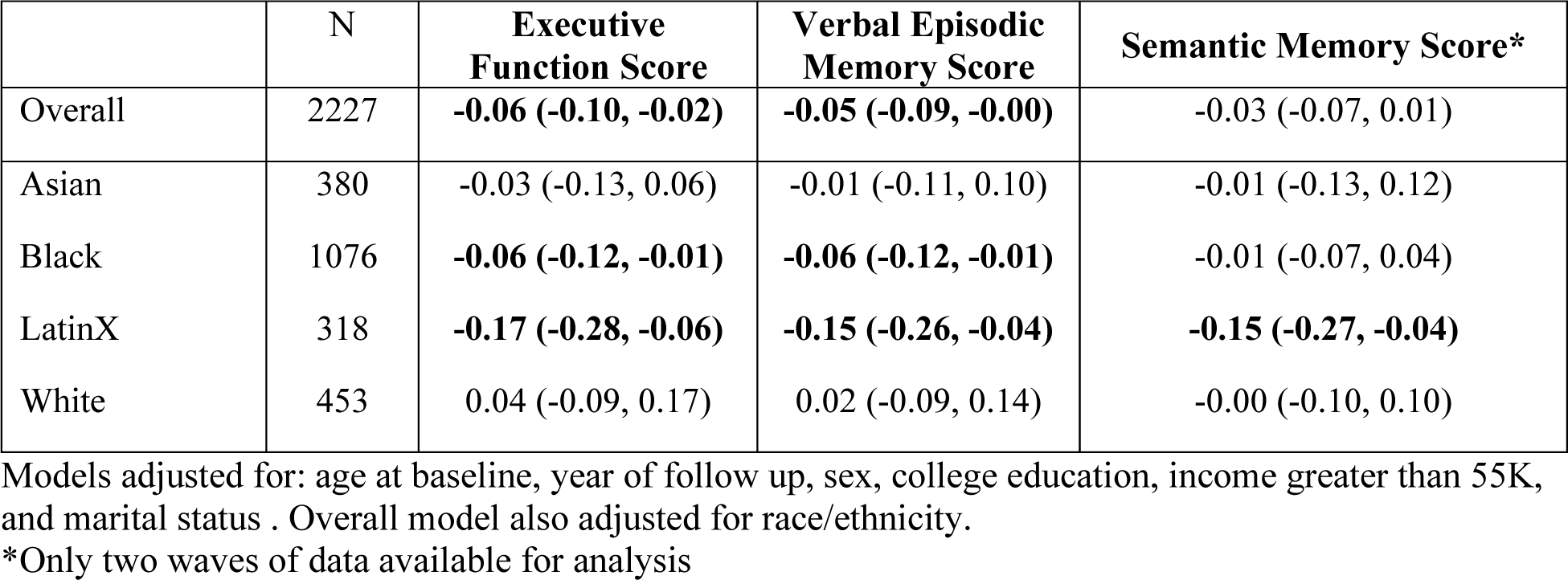
Estimated associations between depressive symptoms and baseline cognitive function among participants in KHANDLE and STAR (N= 2227)

|  | N | **Executive**<br>**Function Score** | **Verbal Episodic**<br>**Memory Score** | **Semantic Memory Score\*** |
| --- | --- | --- | --- | --- |
| Overall | 2227 | **-0.06 (-0.10, -0.02)** | **-0.05 (-0.09, -0.00)** | -0.03 (-0.07, 0.01) |
| Asian | 380 | -0.03 (-0.13, 0.06) | -0.01 (-0.11, 0.10) | -0.01 (-0.13, 0.12) |
| Black | 1076 | **-0.06 (-0.12, -0.01)** | **-0.06 (-0.12, -0.01)** | -0.01 (-0.07, 0.04) |
| LatinX | 318 | **-0.17 (-0.28, -0.06)** | **-0.15 (-0.26, -0.04)** | **-0.15 (-0.27, -0.04)** |
| White | 453 | 0.04 (-0.09, 0.17) | 0.02 (-0.09, 0.14) | -0.00 (-0.10, 0.10) |

Models adjusted for: age at baseline, year of follow up, sex, college education, income greater than 55K, and marital status . Overall model also adjusted for race/ethnicity.
\*Only two waves of data available for analysis

**Table 3.** Estimated associations between depressive symptoms and annual rate of cognitive decline among participants in KHANDLE and STAR (N= 2227)

|  | N | **Executive Function**<br>**Score** | **Verbal Episodic**<br>**Memory Score** | **Semantic Memory**<br>**Score\*** |
| --- | --- | --- | --- | --- |
| Overall | 2227 | -0.01 (-0.02, 0.01) | 0.00 (-0.02, 0.02) | **-0.04 (-0.06, -0.02)** |
| Asian | 380 | 0.00 (-0.02, 0.03) | 0.00 (-0.04, 0.04) | -0.03 (-0.08, 0.01) |
| Black | 1076 | -0.02 (-0.03, 0.00) | **-0.03 (-0.05, -0.00)** | **-0.04 (-0.07, -0.01)** |
| LatinX | 318 | -0.03 (-0.06, 0.01) | 0.02 (-0.03, 0.07) | **-0.10 (-0.15, -0.05)** |
| White | 453 | 0.01 (-0.02, 0.04) | 0.04 (-0.00, 0.09) | 0.00 (-0.04, 0.04) |

Models adjusted for: age at baseline, sex, college education, income greater than 55K, and marital status. Overall model also adjusted for race/ethnicity.
\*Only two waves of data available for analysis.

### Cognitive Decline

In the full sample, we observed that higher depressive symptoms were associated with accelerated decline in semantic memory score (difference in annual rate of change = −0.04 SD, 95%CI: −0.06, −0.02). In the stratified models, among Black participants, we found that more depressive symptoms were associated with accelerated decline in verbal episodic memory (difference in annual rate of change =-0.03 SD, 95%CI: −0.05, −0.00) and semantic memory score (difference in annual rate of change =-0.04 SD, 95%CI: −0.07, −0.01). More depressive symptoms were associated with a rate of decline in semantic memory score that was faster by - 0.10 SD (95%CI: −0.15, −0.05) among LatinX participants, but depressive symptoms were not associated with rate of decline in semantic memory among Asian or White participants. We did not observe associations between depressive symptoms and rate of decline in executive function score in the full sample or in stratified results. Results of the sensitivity analysis using the multiply imputed data showed similar associations between baseline depressive symptoms and accelerated decline (e.g., difference in annual rate of change in semantic memory for the full sample = −0.04 SD, 95%CI: −0.06, −0.02; Table S2). Further, in models including interaction terms of time for all covariates, results were similar (e.g., difference in annual rate of change in semantic memory for the full sample = −0.04 SD, 95%CI: −0.06, −0.02; Table S3). Finally, further adjusting for additional lifestyle covariates and practice setting were somewhat attenuated and confidence intervals were wider (Table S4-S5).

## DISCUSSION

In a diverse sample of adults (ages 50+) followed for up to four years, more depressive symptoms were associated with worse baseline cognitive function as well as faster cognitive decline for Black and LatinX participants in specific cognitive domains over time. Associations differed across cognitive domains and were modified by race and ethnicity. Associations were especially large for LatinX individuals, with effect sizes several times the magnitude as in White respondents.

Our findings on lower depression symptoms among Asian and Black participants, compared to LatinX and White participants are consistent with racial and ethnic differences in depression that have been documented before. For example, Asian American adults, particularly foreign-born Asian Americans, have been reported to have the lowest lifetime prevalence of mental disorders, including depression, compared to Black, LatinX, and White participatns.^24,25^ In addition, according to the National Center for Health Statistics, in 2019, non-Hispanic Asian adults were least likely to experience depressive symptoms compared with Hispanic, non-Hispanic white, and non-Hispanic black adults.^26^ Black individuals have been reported to have lower lifetime and 12-month rates of major depressive disorder compared with White individuals.^27^ The “Black-white depression paradox” describes the lower prevalence of depression among non-Hispanic Black (relative to non-Hispanic white) individuals despite their greater exposure to major life stressors, a phenomenon that remains unexplained.^28^ The combination of higher average levels of depressive symptoms and larger association with cognitive outcomes among LatinX individuals elevates the likely population impact of depressive symptoms on cognitive outcomes among LatinX older adults.

In models including the full sample, depressive symptoms were associated with worse scores in both executive function and episodic memory. This is consistent with previous studies reporting that depression may negatively impact risk of mild cognitive impairment^29^ and neurocognitive functions. We found that associations were particularly marked in a multi-ethnic older sample for cognitive domains that are governed by fronto-subcortical networks, such as executive functions.^30^ Further, our results suggested effect modification of the depression symptoms and baseline cognitive function association by race and ethnicity for some cognitive domains. We observed negative associations between depressive symptoms and cognition (executive function and verbal episodic memory scores) among Black and LatinX participants, but not among Asian or White participants. Our results on higher depressive symptoms associated with lower executive function and verbal episodic memory scores among Black participants are consistent with an analysis that showed that depressive symptoms were associated with worse task-switching, inhibition, and episodic memory among Black participants, and suggested that Black participants may be more vulnerable to negative effects of depression on cognition than White participants.^31^ Our results on higher depressive symptoms associated with lower executive function and verbal episodic memory scores among LatinX participants are consistent with previous work showing that greater depressive symptoms were associated with worse episodic memory among Caribbean Hispanic older adults.^32^ Among Asian participants, prior findings are mixed regarding associations between depressive symptoms and cognition, with a cross-sectional study showing increased prevalence of depressive symptoms in cognitively impaired Chinese American participants,^33^ while a longitudinal study found that more severe depressive symptoms were associated with better cognitive function among patients in China.^34^

With regards to cognitive decline, in models including the full sample, we observed that more depressive symptoms were associated with a faster rate of cognitive decline in semantic memory scores but not in other cognitive domains. Null associations between depressive symptoms and change in cognitive performance across time were also observed in a randomized trial of 445 middle-to-older aged adults designed to compare the effectiveness of internet cognitive behavioral therapy relative to an online attention control.^35^ In our sample, the association between depressive symptoms and cognitive decline was modified by race and ethnicity. More depressive symptoms were associated with a faster rate of cognitive decline in verbal episodic memory among Black participants. This is consistent with a recent finding from the Washington/Hamilton Heights Inwood Columbia Aging Project (WHICAP), a multi-ethnic cohort including White, Black and Hispanic participants, that reported that a higher baseline level of depressive symptoms was associated with a faster decline in episodic memory.^4^ The results on rate of cognitive decline for Black participants despite the lower scores for depression symptoms (compared to LatinX and White participants) could be due to the fact that when depression affects Black participants, it is often untreated and more severe compared to White participants, and therefore, when Black participants report heightened depressive symptoms, they may more significantly influence cognitive functioning than in other groups.^13^ Simultaneously, researchers have observed that Black participants have reduced access to mental health services, and often receive poorer quality care than White participants.^36^ Along these lines, disparities in primary care (e.g., counselling/referrals for counseling, antidepressant medications) persist, with Black and LatinX participants having less access to care than White participants.^9^ Thus, higher depression symptoms in Black participants might be indicative of untreated depression and seemed associated with a steeper decline in cognition. More depressive symptoms were associated with a faster rate of cognitive decline in semantic memory scores among Black and LatinX participants, although these results should be interpreted with caution due to limited time-points available. Finally, we did not observe an association between depressive symptoms and rate of cognitive decline in analyses restricted to Asian or White participants. Reasons for this, may include the low prevalence of depressive symptoms among Asian and White participants, resulting in imprecise estimates with wide confidence intervals for these groups.^37^

Several limitations in this study should be noted. First, disease mechanisms common to depression and cognitive health may explain the associations observed. It remains controversial whether depressive symptoms increase risk of dementia, are an early symptom of neurodegeneration, or are a reaction to early cognitive deficit. To account for this, we used longitudinal data including 3 repeated measurements of cognitive performance to evaluate trajectories of cognitive decline. We note that prior studies with much longer follow-up have helped establish temporal order; for example a study from Denmark demonstrated that depression diagnoses predicted incident dementia over 20 years later.^4^ The major contribution in the current study is to demonstrate the relevance of depressive symptoms for Black and LatinX adults, who were not represented in much prior work. However, we only had two assessments of semantic memory outcome due to the shift to phone administration of KHANDLE visits during the COVID-19 pandemic, and thus our estimates of semantic memory decline over time have a shorter follow up and are less precise. Second, participants of Latinx/Hispanic origin include ethnic/regional subgroups (e.g. Mexicans, Dominicans, Puerto Ricans, etc.) with important intergroup differences; Asian Americans are similarly heterogeneous. However, we were under powered to assess LatinX or Asian subgroup differences in the association between depression symptoms and cognitive health. Future more larger-scale, racially and ethnically diverse studies are needed. Despite these limitations, this study has notable strengths. The use of longitudinal data in the KHANDLE and STAR cohorts allowed us to examine prospective associations between depression and cognitive decline. The study was conducted in an exceptionally diverse cohort, with rich data on covariates that allowed us to adjust for important confounders. We also had a well-validated neuropsychological assessment scales enabling us to examine domain-specific associations.

Among participants in KHANDLE and STAR, greater depressive symptoms were associated with worse cognitive health and potentially faster cognitive decline among certain cognitive domains and race and ethnicity groups. Our findings provide support for the hypothesis that Black and LatinX participants are more susceptible to the negative effects of depressive symptoms on cognitive health.

## Data Availability

Data used in the present study can be accessed via a data use request at https://sites.google.com/g.ucla.edu/khandle-study-site/home.

## Acknowledgements

The authors thank the staff and participants of the KHANDLE, and STAR studies for their important contributions. This study is supported by the National Institute on Aging (NIA) R01AG052132, RF1AG050782. MPJ is supported by 4R00AG066949-03.

