## Supplementary Material for "Racial and Ethnic Differences in the Association between Depressive Symptoms and Cognitive Outcomes in Older Adults: Findings from KHANDLE and STAR"


- Figure S1. Data Flow Chart
- Table S1: Estimated associations between depressive symptoms and baseline cognitive function using multiply imputed data
- Table S2: Estimated associations between depressive symptoms and annual rate of cognitive decline using multiply imputed data
- Table S3: Estimated associations between depressive symptoms and annual rate of cognitive decline, including time interactions for all covariates
- Methods S1: Description of additional covariates
- Table S4: Estimated associations between depressive symptoms and baseline cognitive function after further adjustment for smoking, heavy drinking, daily physical activity, daily socialization, self-reported health, and practice setting
- Table S5: Estimated associations between depressive symptoms and annual rate of cognitive decline after further adjustment for smoking, heavy drinking, daily physical activity, daily socialization, self-reported health, and practice setting

Figure S1. Data Flow Chart

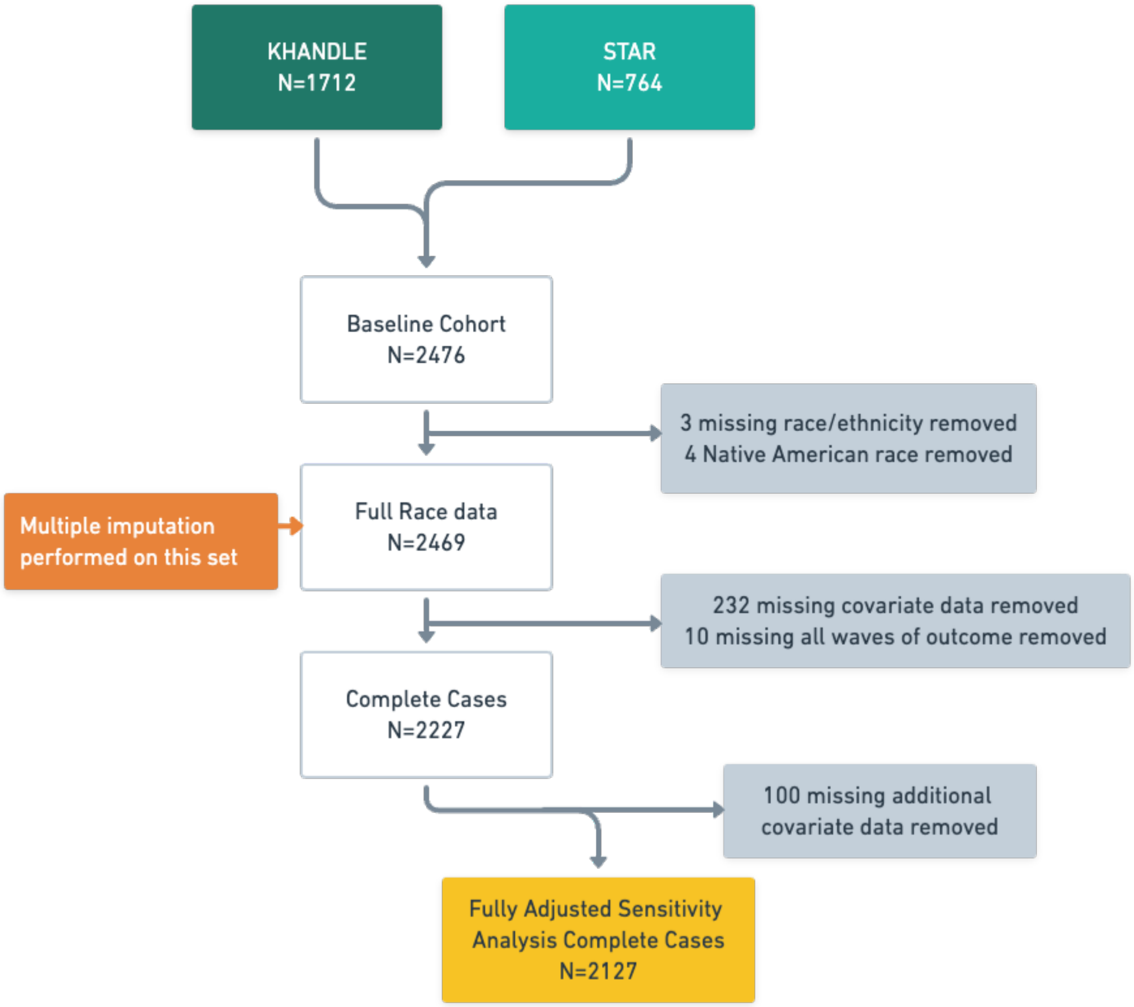


NOTE: Variables used to define the complete cases included: age at baseline, sex, education, income, and marital status. Additional covariates considered to create complete case set for the sensitivity analysis included: smoking status, heavy drinking, daily physical activity, daily socializing, self-reported health, and practice setting

Table S1. Estimated associations between depressive symptoms and baseline cognitive function using multiply imputed data. KHANDLE and STAR (N= 2469)

| | N | **Executive Function Score** | **Verbal Episodic Memory Score** | **Semantic Memory Score*** |
|---|---|---|---|---|
| Overall | 2469 | **-0.07 (-0.11, -0.03)** | **-0.06 (-0.10, -0.02)** | -0.04 (-0.08, 0.00) |
| Asian | 415 | -0.04 (-0.14, 0.05) | 0.00 (-0.10, 0.11) | -0.01 (-0.13, 0.11) |
| Black | 1197 | **-0.07 (-0.13, -0.02)** | **-0.08 (-0.13, -0.02)** | -0.02 (-0.08, 0.03) |
| LatinX | 355 | **-0.15 (-0.25, -0.04)** | **-0.11 (-0.22, -0.00)** | **-0.16 (-0.27, -0.04)** |
| White | 502 | -0.03 (-0.15, 0.09) | -0.02 (-0.13, 0.09) | -0.02 (-0.13, 0.09) |

Models adjusted for: age at baseline, year of follow up, sex, college education, income greater than 55K, and marital status Overall model also adjusted for race/ethnicity.
*Only two waves of data available for analysis

Table S2. Estimated associations between depressive symptoms and annual rate of cognitive decline using multiply imputed data. KHANDLE and STAR (N= 2469)

| | N | **Executive Function Score** | **Verbal Episodic Memory Score** | **Semantic Memory Score*** |
|---|---|---|---|---|
| Overall | 2469 | -0.01 (-0.02, 0.00) | 0.00 (-0.02, 0.02) | **-0.04 (-0.06, -0.02)** |
| Asian | 415 | 0.00 (-0.02, 0.03) | 0.00 (-0.04, 0.04) | -0.04 (-0.08, 0.00) |
| Black | 1197 | -0.01 (-0.03, 0.01) | -0.02 (-0.04, 0.01) | **-0.04 (-0.07, -0.02)** |
| LatinX | 355 | -0.01 (-0.04, 0.02) | 0.02 (-0.03, 0.07) | **-0.08 (-0.12, -0.04)** |
| White | 502 | 0.00 (-0.03, 0.03) | 0.02 (-0.02, 0.06) | -0.01 (-0.05, 0.03) |

Models adjusted for: age at baseline, sex, college education, income greater than 55K, and marital status. Overall model also adjusted for race/ethnicity.
*Only two waves of data available for analysis

Table S3. Estimated associations between depressive symptoms and annual rate of cognitive decline, including time interactions for all covariates. KHANDLE and STAR (N= 2237)

| | N | **Executive Function Score** | **Verbal Episodic Memory Score** | **Semantic Memory Score*** |
|---|---|---|---|---|
| Overall | 2227 | -0.01 (-0.02, 0.01) | 0.00 (-0.02, 0.02) | **-0.04 (-0.06, -0.02)** |
| Asian | 380 | 0.01 (-0.02, 0.03) | 0.00 (-0.04, 0.04) | -0.04 (-0.08, 0.01) |
| Black | 1076 | -0.02 (-0.03, 0.00) | **-0.03 (-0.05, -0.00)** | **-0.04 (-0.07, -0.01)** |
| LatinX | 318 | -0.02 (-0.06, 0.01) | 0.01 (-0.04, 0.07) | **-0.10 (-0.15, -0.05)** |
| White | 453 | 0.02 (-0.01, 0.05) | 0.04 (-0.00, 0.09) | 0.00 (-0.04, 0.04) |

Models adjusted for: age at baseline, sex, college education, income greater than 55K, and marital status. All covariates were included with interactions for years since baseline.
*Only two waves of data available for analysis

Methods S1

Sensitivity analyses in Tables S4-S5 are an extension of complete case analyses reported in Tables 2-3 from the main paper, further adjusting for smoking, heavy drinking, daily physical activity, daily socialization, self-reported health, and practice setting. All additional adjustment variables are binary.

**Smoking** is defined as ever reported smoking vs. never reported smoking.

**Heavy drinking** is defined as drinking every day in the past three months or having four or more drinks when drinking vs. not.

**Daily physical activity** is defined as yes if participants report any of the following activities with a daily or almost every day frequency: Light house or yard work (tidying, dusting, sweeping, laundry, gardening, etc); Light exercise or sports (walking, dancing, softball, bowling, etc); Heavy, work-related demands (carrying, lifting, moving, etc); Vigorous exercise or sports (cycling, jogging, swimming laps, tennis, etc); Vigorous house or yard work (vacuuming, mopping, mowing lawn, etc).

**Daily socialization** is defined as yes if participants reported “Socializing/talking with friends and family” as “Every day, or almost every day.”

**Self-reported health** was defined as excellent or very good vs. not.

**Practice setting** is a three category variable that records the setting in which the interview at each wave took place. Options include: home, office/clinic, and phone.

Table S4. Estimated associations between depressive symptoms and baseline cognitive function, after further adjustment for smoking, heavy drinking, daily physical activity, daily socialization, self-reported health, and practice setting. KHANDLE and STAR (N= 2127)

| | N | **Executive Function Score** | **Verbal Episodic Memory Score** | **Semantic Memory Score*** |
|---|---|---|---|---|
| Overall | 2138 | -0.03 (-0.06, 0.00) | -0.03 (-0.07, 0.01) | -0.03 (-0.07, 0.01) |
| Asian | 358 | -0.02 (-0.10, 0.06) | 0.00 (-0.10, 0.10) | -0.03 (-0.15, 0.08) |
| Black | 1053 | -0.04 (-0.08, 0.00) | **-0.06 (-0.11, -0.01)** | -0.02 (-0.07, 0.03) |
| LatinX | 290 | **-0.10 (-0.19, -0.01)** | **-0.11 (-0.22, -0.01)** | -0.09 (-0.20, 0.02) |
| White | 426 | 0.06 (-0.04, 0.16) | 0.06 (-0.04, 0.16) | 0.00 (-0.10, 0.09) |

Models adjusted for: age at baseline, year of follow up, sex, college education, income greater than 55K, marital status, smoking status, heavy drinking, daily physical activity, daily socializing, self-reported health, and practice setting
*Only two waves of data available for analysis

Table S5. Estimated associations between depressive symptoms and annual rate of cognitive decline after further adjustment for smoking, heavy drinking, daily physical activity, daily socialization, self-reported health, and practice setting. KHANDLE and STAR (N= 2127)

| | N | **Executive Function Score** | **Verbal Episodic Memory Score** | **Semantic Memory Score*** |
|---|---|---|---|---|
| Overall | 2138 | -0.01 (-0.02, 0.00) | 0.00 (-0.02, 0.01) | **-0.03 (-0.05, -0.01)** |
| Asian | 358 | 0.00 (-0.02, 0.02) | 0.00 (-0.04, 0.04) | -0.03 (-0.07, 0.01) |
| Black | 1053 | -0.01 (-0.02, 0.00) | -0.03 (-0.05, 0.00) | **-0.03 (-0.06, -0.01)** |
| LatinX | 290 | -0.02 (-0.05, 0.00) | 0.00 (-0.05, 0.05) | **-0.08 (-0.13, -0.04)** |
| White | 426 | 0.01 (-0.02, 0.03) | 0.03 (-0.01, 0.07) | 0.01 (-0.02, 0.05) |

Models adjusted for: age at baseline, sex, college education, income greater than 55K, marital status, smoking status, heavy drinking, daily physical activity, daily socializing, self-reported health, and practice setting.
*Only two waves of data available for analysis
